# Warming, thermal variability, and the 96% decline in childhood respiratory-infection mortality in China: a national time-series analysis of the Global Burden of Disease Study 2021 and the C-LSAT high-resolution climate dataset

**DOI:** 10.64898/2026.08.31.26361879

**Authors:** Deze Lia, Hao Chen, Xiaotong Wang, Chen Shen

**Affiliations:** Beijing Pediatric Research Institute, Beijing Children’s Hospital, Capital Medical University, Beijing, China

**Keywords:** temperature variability, climate change, child health, respiratory infections, minimum mortality temperature, China, Global Burden of Disease

## Abstract

**Background:** Childhood respiratory mortality in China has fallen by over 90% in three decades alongside sustained national warming, yet national long-run evidence on temperature and child respiratory mortality is lacking.

**Methods:** We linked Global Burden of Disease (GBD) 2021 mortality estimates (rates use a fixed 2022 population denominator, so rate trends equal death-count trends) for China — lower respiratory infections (LRI), ages 0–19, and asthma, ages 0–24, 1990–2021 — with C-LSAT 0.5° gridded temperature data (1990–2019), aggregated nationally and to five climate zones. Four annual indicators (mean temperature, diurnal temperature range, seasonal amplitude, interannual variability) entered regressions of log mortality rates with Newey–West standard errors. A bootstrapped (500 resamples) quadratic model probed the minimum mortality temperature (MMT), with PM2.5-adjusted analyses and future-exposure, permutation, and detrended falsification tests. Changes in China’s national thermal regime for 2041–2060 were additionally projected under three CMIP6 SSP scenarios (SSP1-2.6, SSP2-4.5 and SSP5-8.5; WorldClim v2.1 1970–2000 baseline, MRI-ESM2-0) using a matched-cell delta method.

**Results:** LRI deaths fell by 96.3% (330,194 in 1990 to 12,098 in 2021; 95% uncertainty interval 9,669–14,891) and asthma deaths by 94.9% (3,287 to 167), while mean temperature rose 0.364 °C per decade and diurnal temperature range narrowed 0.092 °C per decade. Baseline coefficients were large (mean temperature −1.696, SE 0.174; diurnal temperature range +2.408, SE 0.336; seasonal amplitude −0.162, SE 0.082; interannual variability +2.924, SE 1.514, per 1 °C in log rate), but the future-exposure test failed and detrending nullified every coefficient: the associations are trend-level, and short-cycle causal effects are not identifiable. Nor was the national MMT identifiable — observed temperature support spans only 6.66–8.13 °C, and the nominal turning point of 35.84 °C is an extrapolation artifact (quadratic term p = 0.963). Within the observed range, warming and declining mortality moved in the same direction. A CMIP6 scenario projection (WorldClim v2.1, MRI-ESM2-0, matched-cell national means) indicates a further +2.19 to +3.02 °C of national warming by 2041–2060, with little change in diurnal temperature range or seasonal amplitude.

**Conclusions:** The 96% decline in childhood respiratory mortality cannot be attributed to warming. China sits on the low-temperature side of the optimum, and the marginal direction of future warming requires stronger designs to establish. The falsification framework offers a discipline for climate–health inference in China. Bias-corrected scenario projections place China’s 2050s mean temperature (approximately 9.4–10.3 °C on the C-LSAT scale) still far below multi-country optima (14.7–19.8 °C), noting that an annual-mean construct is not fully comparable with daily-scale MMT estimates.

## 1. Introduction

Ambient temperature is a well-established determinant of human mortality. In a 13-country analysis, 7.71% of deaths were attributable to non-optimal temperatures, with cold (7.29%) outweighing heat (0.42%) by roughly seventeen-fold.^1^ A global assessment estimated 5,083,173 temperature-related deaths per year during 2000–2019 (9.43% of all deaths), again dominated by cold (8.52% versus 0.91%).^2^ The temperature at which mortality is lowest — the minimum mortality temperature (MMT) — varies with geography and local climate, and its estimation carries substantial uncertainty.^3^ ^1^ ^4^ Beyond the mean level, thermal variability itself is hazardous: diurnal temperature range (DTR) has been linked to mortality in multi-country studies and systematic reviews, with an estimated attributable fraction of 2.5%.^5^ ^6^ ^7^ In Shanghai, each 1 °C increase in DTR raised respiratory mortality by 1.29% on cold days.^8^

Children may be especially susceptible, yet paediatric evidence is thin. Infants under one year appear most vulnerable to heat, and temperature effects in children operate mainly through infectious diseases, including respiratory infections.^9^ A 5 °C increase in DTR has been associated with a 31% increase in childhood asthma emergency visits,^10^ and exposure is rising: infants worldwide experienced an average of 13.8 heatwave days in 2023, 8.2 days above the historical baseline.^11^ However, this evidence is almost entirely city-level and short-term;^12^ national, long-run analyses of temperature and child respiratory mortality are lacking, particularly for China.

This gap is striking given China’s recent history. Between 1990 and 2021, deaths from lower respiratory infections (LRI) among Chinese children aged 0–19 years fell by 96.3%, from 330,194 to 12,098, and asthma deaths among ages 0–24 years fell by 94.9%, from 3,287 to 167.^13,14^ This decline — among the largest reductions in child mortality on record — unfolded in parallel with sustained national warming. We therefore pose three questions. First, how is the three-decade decline in childhood respiratory mortality related to concurrent warming? Second, does thermal variability — operationalised as DTR, seasonal amplitude, and interannual fluctuation — show associations independent of mean temperature? Third, where does China’s childhood MMT lie, and is it identifiable from the observed climate range?

To address these questions, we linked the complete national GBD 2021 mortality series for China (1990–2021) with the C-LSAT 0.5° high-resolution land surface temperature and DTR datasets.^15^ Our contributions are threefold: (i) the first national-scale coupling of the full GBD childhood respiratory mortality series with high-resolution Chinese climate data; (ii) a four-dimensional decomposition of thermal exposure — mean, DTR, seasonal amplitude, and interannual variability — with adjustment for ambient fine particulate matter (PM2.5);^16^ and (iii) an explicit falsification framework (future-exposure, permutation, and detrended specifications) with a formal identifiability assessment of the national MMT, in which every turning-point estimate is reported against the observed temperature support (6.66–8.13 °C), so that extrapolated values are labelled as such rather than read as identified optima.

## 2. Methods

### 2.1 Data sources

Mortality outcomes were drawn from the Global Burden of Disease (GBD) 2021 study for China, 1990–2021.^13^ ^14^ The primary outcome was deaths from lower respiratory infections (LRI; GBD cause 322) among children aged 0–19 years, obtained by summing the four constituent age groups (<5, 5–9, 10–14, and 15–19 years); the 20–24-year group was never combined into the 0– 19 total. The secondary outcome was asthma deaths (GBD cause 515) among ages 0–24 years, the narrowest paediatric age band available for this cause. Climate exposures came from the C-LSAT high-resolution land surface air temperature dataset (HRv1) and its paired diurnal temperature range product (C-LDTR), which provide gridded monthly temperature and diurnal temperature range at 0.5° resolution over 1901–2023; the regression panel used 1990–2019 to avoid the COVID-19 perturbation years, which disrupted both mortality ascertainment and healthcare use; the 2020–2021 mortality years were retained for trend description only.^15^ As a time-varying covariate we used the GBD 2023 summary exposure value (SEV) for ambient particulate matter (PM2.5) for China, 1990–2023.^16^ We note a version difference across inputs: mortality outcomes are from GBD 2021, whereas the PM2.5 SEV series is from GBD 2023; the two rounds use partly different estimation pipelines, and we therefore treat the PM2.5-adjusted model as a sensitivity analysis rather than the primary specification.

### 2.2 Exposure construction and climate zones

Gridded C-LSAT values were aggregated to 33 province-level units (GADM 4.1 boundaries) and then to the national level using land-area weights. Four annual indicators were constructed per unit: (i) *mean_temp*, the annual mean temperature; (ii) *dtr_mean*, the annual mean of daily diurnal temperature range; (iii) *season_amp*, the within-year seasonal amplitude, defined as the difference between the warmest and coldest monthly mean temperatures; and (iv) *interannual_sd*, the standard deviation of annual mean temperature over a trailing 10-year window, capturing year-to-year thermal variability. For zone-stratified description, provinces were grouped into five climate zones: cold temperate (Heilongjiang, Jilin, Liaoning, NeiMongol, XinjiangUygur), mid temperate (Beijing, Tianjin, Hebei, Shanxi, Shandong, Henan, Shaanxi, Gansu, NingxiaHui), warm temperate (Shanghai, Jiangsu, Anhui, Zhejiang, Hubei, Hunan, Jiangxi, Sichuan, Chongqing, Guizhou), subtropical (Fujian, Guangdong, Guangxi, Hainan, Yunnan, HongKong, Macau), and plateau (Qinghai, Xizang). Because provincial paediatric population time series were unavailable, national aggregation used area rather than population weights; this over-weights large western provinces relative to the populous east and is revisited as a limitation.

### 2.3 Outcomes and rates

Both outcomes were expressed as rates per 100,000 using a single fixed denominator: the GLOBOCAN 2022 estimate of the Chinese population aged 0–19 years (317,671,474), a level consistent with the GBD 2023 demographic estimates for China.^17^ A fixed denominator isolates the mortality-numerator dynamics and avoids importing population-age-structure change into the trend, at the cost that the resulting series is a mortality index rather than an age-standardised rate. For asthma, GBD deaths span ages 0–24 years while the denominator covers ages 0–19 years; this numerator–denominator mismatch is deliberate under the fixed-denominator convention and affects levels, not within-series relative change, which is the quantity entering all regressions.

### 2.4 Statistical models

The primary specification was an ordinary least squares regression of the natural logarithm of the national mortality rate on the four climate indicators jointly:

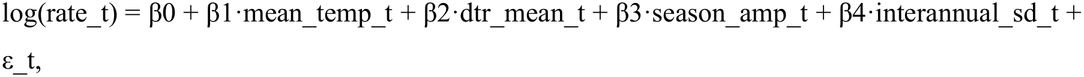

estimated on n = 30 annual observations (1990–2019). Standard errors were computed with Newey–West heteroskedasticity- and autocorrelation-consistent (HAC) covariance with a maximum lag of 3 years, given the short, strongly autocorrelated series. Coefficients are interpreted as the change in log mortality rate per +1 °C of the indicator. Two sensitivity analyses were pre-specified: (i) adding the national PM2.5 SEV as a covariate; and (ii) a linearly detrended model in which the log rate and all regressors were residualised on calendar year, removing shared secular trends.

### 2.5 Minimum mortality temperature

To probe a national minimum mortality temperature (MMT), we fitted a quadratic in mean_temp (log rate on mean_temp and mean_temp²) and located the turning point at −β1/2β2.^1^ ^18^ Uncertainty was quantified with 500 bootstrap resamples over years; draws yielding a non-positive quadratic coefficient (no minimum) were discarded from the interval, and the proportion of such draws was reported. We applied a strict observed-support rule: any turning point must be reported together with the observed range of the exposure, and any estimate lying outside that range is explicitly labelled an extrapolation rather than an identified MMT.

### 2.6 Falsification suite

Three falsification tests assessed whether baseline associations reflect short-run causal structure or shared trends: (i) a future-exposure test replacing concurrent exposures with their t+1 and t+2 leads, which should attenuate a genuinely causal concurrent effect; (ii) 200 permutations of calendar years to generate a null distribution for the coefficients; and (iii) the detrended model described above. Failure of the future-exposure test, or null effects after detrending, was taken as evidence against a short-run causal interpretation.

### 2.7 Reporting, ethics, and software

The study used publicly available, de-identified, aggregated estimates and required no ethics approval or consent. Analyses were conducted in Python (statsmodels for HAC estimation; NumPy/pandas for data processing), and all aggregation and modelling code is available from the corresponding author on reasonable request.

### 2.8 Scenario projection of China’s future thermal regime

As forward-looking context, we projected the change in China’s national thermal regime between the WorldClim version 2.1 1970–2000 baseline climatology and CMIP6-downscaled projections for 2041–2060 (10 arc-minute resolution, monthly minimum and maximum temperature; single general circulation model, MRI-ESM2-0)^21^ under SSP1-2.6, SSP2-4.5 and SSP5-8.5. National means were computed over the 34,111 grid cells covered by China’s Natural Earth polygon and valid in the future land mask, with baseline and future averages taken over identical cells (matched-cell delta method). Annual mean temperature, mean diurnal temperature range and seasonal amplitude were derived as in Section 2.2, except that grid cells were unweighted rather than land-area weighted. These are scenario-conditional climatological changes and are reported as exposure context only; no mortality projection is attempted.

## 3. Results

### 3.1 Thirty-year trends in childhood respiratory mortality and climate

the corresponding mortality rate fell from 103.9 to 3.81 per 100,000 (Table 1; rates use the fixed 2022 population denominator, so rate trends equal death-count trends).

**Table 1.**
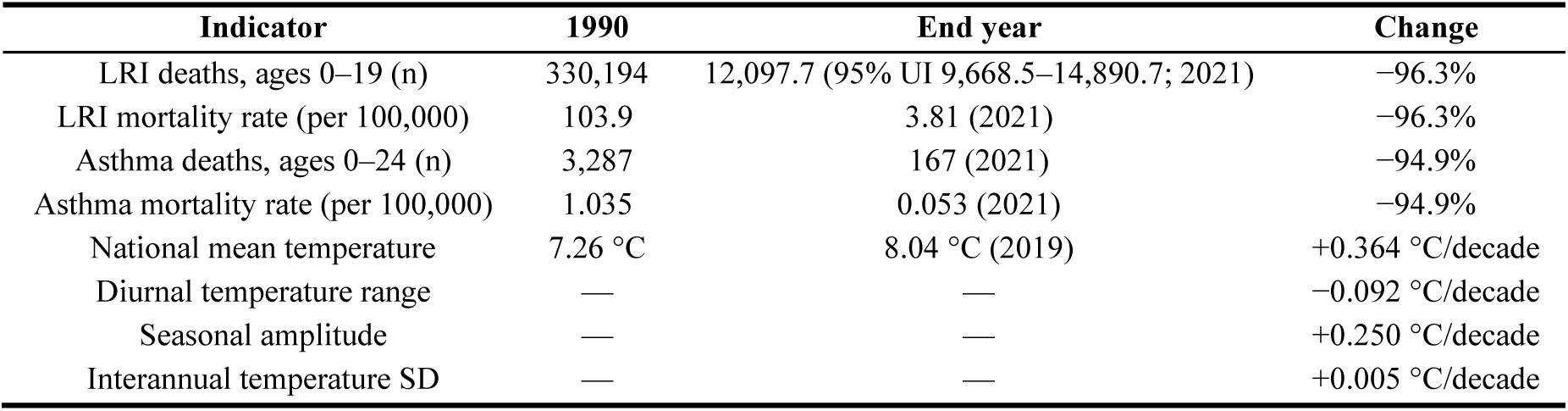
Trend anchors for childhood respiratory mortality (1990–2021) and national climate indicators (1990–2019), China.

| Indicator | 1990 | End year | Change |
|---|---|---|---|
| LRI deaths, ages 0–19 (n) | 330,194 | 12,097.7 (95% UI 9,668.5–14,890.7; 2021) | −96.3% |
| LRI mortality rate (per 100,000) | 103.9 | 3.81 (2021) | −96.3% |
| Asthma deaths, ages 0–24 (n) | 3,287 | 167 (2021) | −94.9% |
| Asthma mortality rate (per 100,000) | 1.035 | 0.053 (2021) | −94.9% |
| National mean temperature | 7.26 °C | 8.04 °C (2019) | +0.364 °C/decade |
| Diurnal temperature range | — | — | −0.092 °C/decade |
| Seasonal amplitude | — | — | +0.250 °C/decade |
| Interannual temperature SD | — | — | +0.005 °C/decade |

The climate series moved concurrently but far more modestly (Figure 1; Table 1). Over 1990– 2019, the area-weighted national mean temperature rose by 0.364 °C per decade, from 7.26 °C in 1990 to 8.04 °C in 2019. Mean diurnal temperature range (DTR) declined by 0.092 °C per decade, whereas seasonal amplitude increased by 0.250 °C per decade. Interannual temperature variability was essentially stable, changing by only +0.005 °C per decade. Thus, the observation window pairs a near-total decline in childhood respiratory mortality with gradual warming, narrowing DTR, widening seasonal amplitude, and unchanged year-to-year variability.

**Figure 1.**
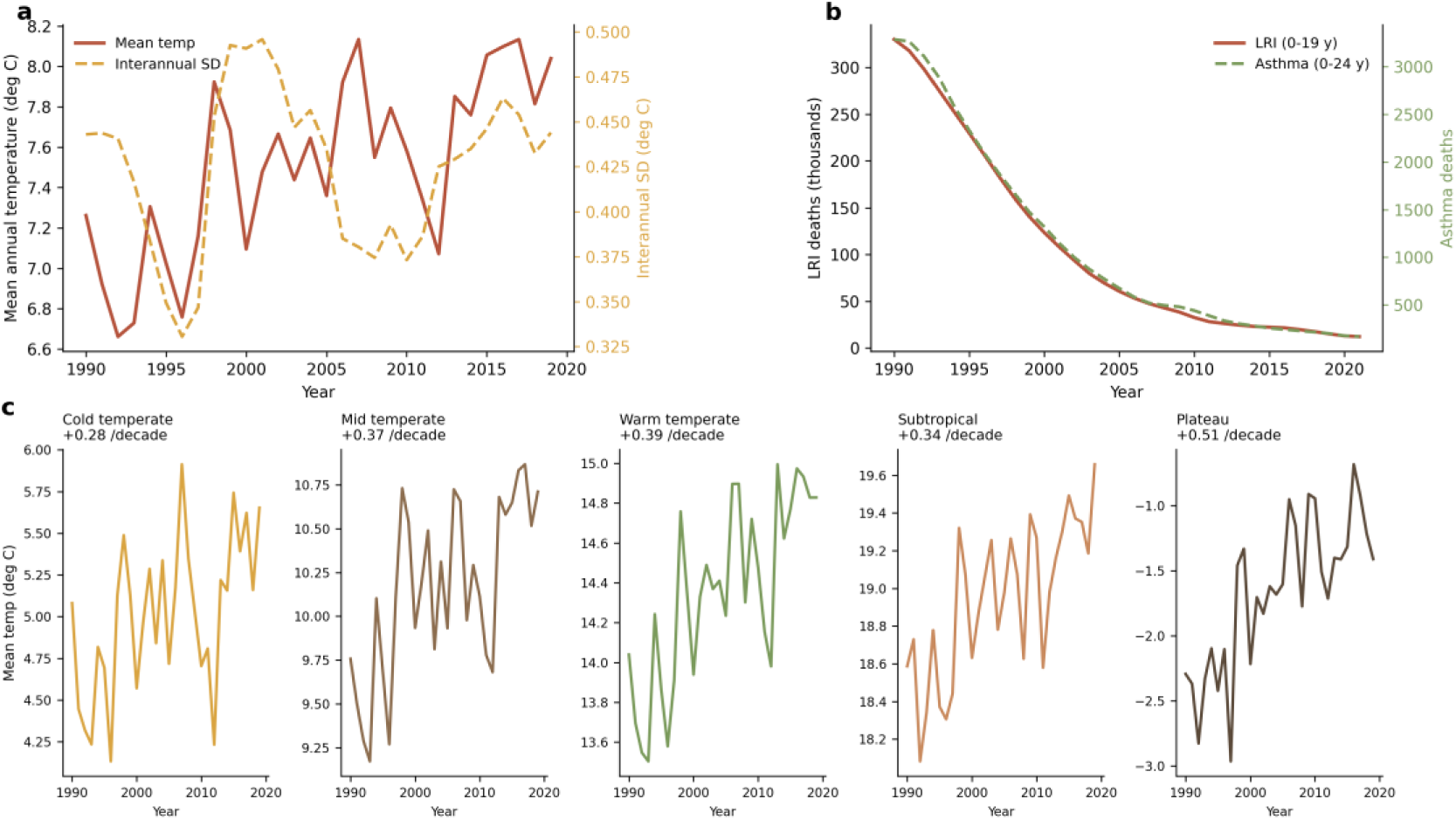
(a) National mean annual temperature (solid) and interannual variability (dashed), 1990–2019 (warming 0.364 °C/decade); (b) childhood LRI deaths (ages 0–19; −96.3%) and asthma deaths (ages 0–24; −94.9%), 1990–2021; (c) mean-temperature trajectories by climate zone (small multiples; per-decade trend shown).

### 3.2 Baseline associations between climate indicators and mortality

Figure 2 summarizes the baseline Newey–West regressions of the log mortality rate on the four climate indicators (n = 30 years, 1990–2019); coefficients denote the change in log mortality per 1 °C increase in each indicator (Table 2). Annual mean temperature was strongly and negatively associated with both outcomes: −1.696 (SE 0.174, p < 0.001) for LRI and −1.600 (SE 0.161, p < 0.001) for asthma. DTR was positively associated with mortality (LRI: +2.408, SE 0.336, p < 0.001; asthma: +2.210, SE 0.294, p < 0.001). Seasonal amplitude showed a weaker negative association (LRI: −0.162, SE 0.082, p = 0.047; asthma: −0.137, SE 0.072, p = 0.056), and interannual variability a weaker positive one (LRI: +2.924, SE 1.514, p = 0.054; asthma: +2.525, SE 1.348, p = 0.061). Bivariate correlations confirmed the direction of the mean-temperature (r = −0.72), DTR (r = +0.50), and seasonal-amplitude (r = −0.23) coefficients; the interannual-variability coefficient reversed sign relative to its weak bivariate correlation (r = −0.09), a sign flip attributable to collinearity among the climate regressors, and is therefore interpreted cautiously.

**Figure 2.**
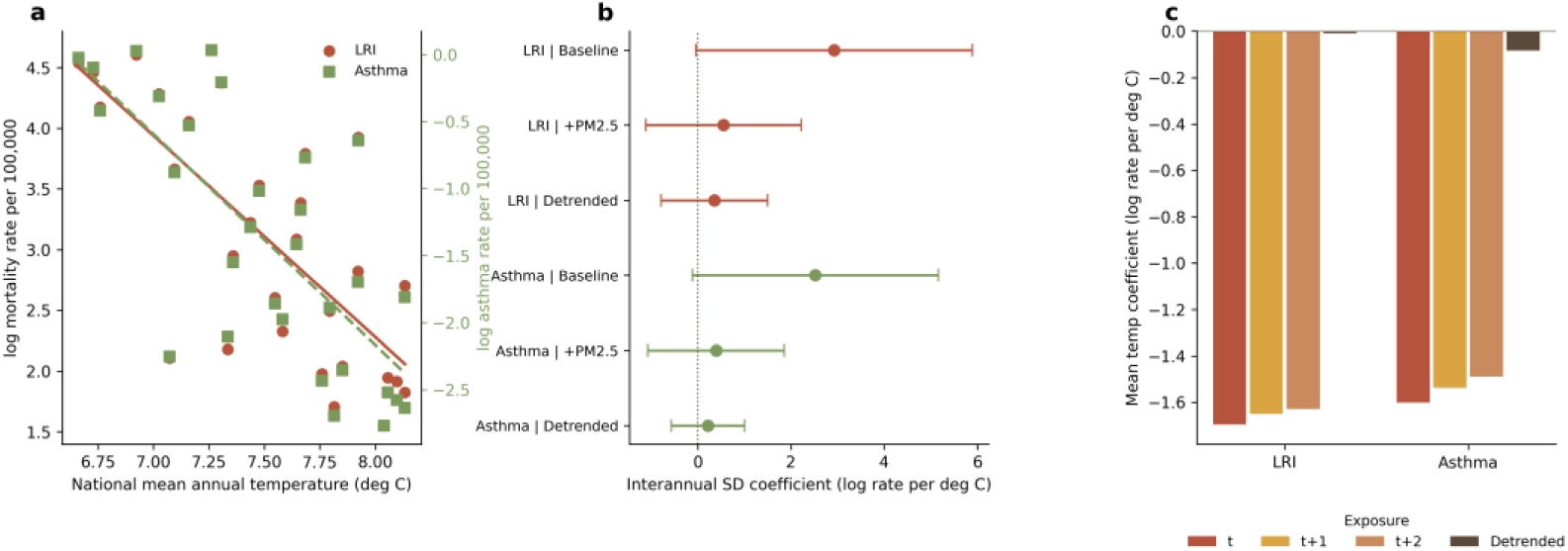
(a) National mean temperature versus log LRI and log asthma mortality rates, 1990–2019, with linear fits; (b) interannual-SD coefficients for LRI (2.92, 95% CI −0.04 to 5.89) and asthma (2.52, −0.12 to 5.17) under baseline, PM2.5-adjusted and detrended specifications; (c) falsification controls: contemporaneous versus future-exposure (t+1, t+2) mean-temperature coefficients and the detrended null.

**Table 2.**
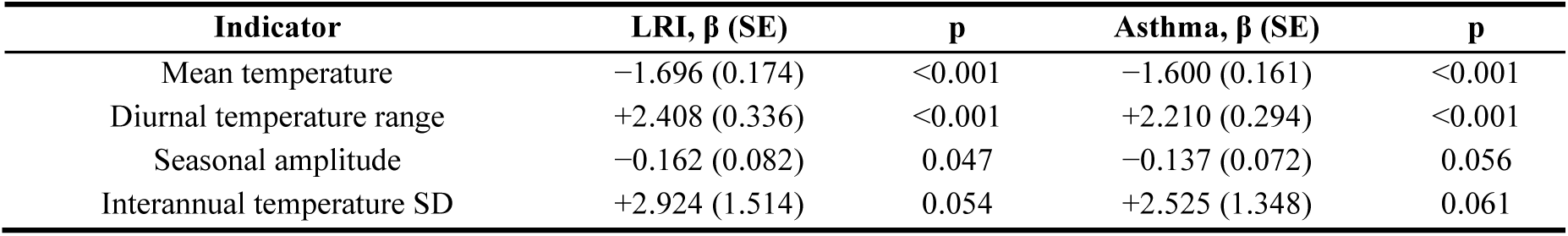
Baseline model coefficients (log mortality rate per 1 °C), Newey–West HAC (lag = 3), n = 30, 1990–2019.

| **Indicator** | **LRI, β (SE)** | **p** | **Asthma, β (SE)** | **p** |
|---|---|---|---|---|
| Mean temperature | −1.696 (0.174) | <0.001 | −1.600 (0.161) | <0.001 |
| Diurnal temperature range | +2.408 (0.336) | <0.001 | +2.210 (0.294) | <0.001 |
| Seasonal amplitude | −0.162 (0.082) | 0.047 | −0.137 (0.072) | 0.056 |
| Interannual temperature SD | +2.924 (1.514) | 0.054 | +2.525 (1.348) | 0.061 |

### 3.3 Adjustment for PM2.5 and detrended models

Adding the PM2.5 summary exposure value to the model attenuated the mean-temperature coefficient by roughly three quarters, to −0.419 (SE 0.140, p = 0.003) for LRI and −0.453 (SE 0.145, p = 0.002) for asthma, although it remained statistically significant. The seasonal-amplitude coefficient also remained negative and significant (−0.077, SE 0.032, p = 0.015 for LRI; −0.061, SE 0.029, p = 0.037 for asthma). In contrast, the DTR and interannual-variability coefficients lost all significance after PM2.5 adjustment (DTR: p = 0.728 and p = 0.625; interannual SD: p = 0.515 and p = 0.597, for LRI and asthma respectively). PM2.5 itself entered with a negative coefficient (−0.070, SE 0.008, p < 0.001 for LRI; −0.063, SE 0.008, p < 0.001 for asthma), co-moving with the long-run mortality decline rather than capturing a short-run effect. In the linearly detrended specification, every climate coefficient collapsed toward zero, with magnitudes shrinking by an order of magnitude and no coefficient approaching significance (all p > 0.24).

### 3.4 The minimum-mortality temperature is not identifiable in China

Figure 3 presents the quadratic temperature–mortality curves. For LRI, the quadratic term was small and non-significant (b2 = 0.029, p = 0.963); the implied turning point was 35.84 °C, with a bootstrap 95% confidence interval of 7.92–46.50 °C, and only 248 of 500 bootstrap resamples yielded a positive quadratic term—that is, an interior minimum existed in fewer than half of resamples. For asthma, the quadratic term was negative and non-significant (b2 = −0.045, p = 0.935), so no minimum-mortality temperature exists. Critically, the observed support of the national annual mean temperature spans only 6.66–8.13 °C across the 30 study years; the LRI point estimate of 35.84 °C lies far outside this support and is an extrapolation artifact of the quadratic form. We therefore judge the national childhood minimum-mortality temperature (MMT) to be not identifiable from this series. Within the observed support, log mortality declined monotonically with mean temperature for both outcomes. China’s national mean temperature (approximately 7.5 °C) sits on the low-temperature side of the minimum-mortality temperatures typically estimated in multi-country studies, so warming and declining mortality move together within the observed range—without implying that further warming would remain beneficial.

**Figure 3.**
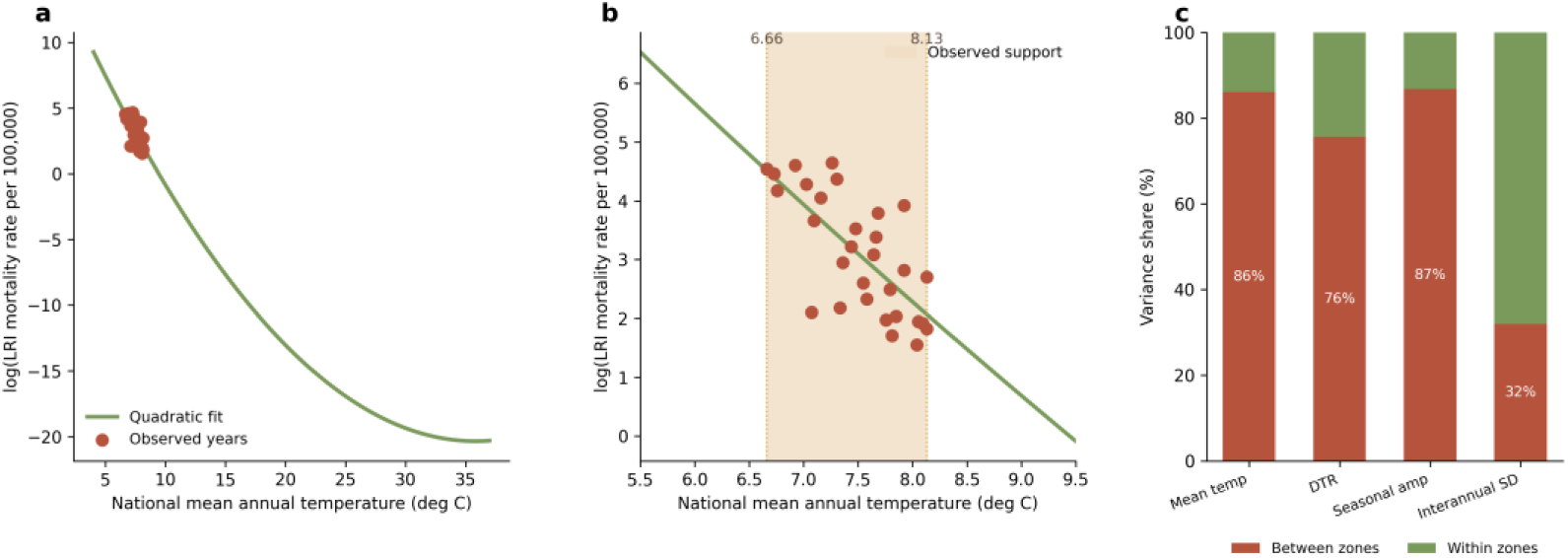
(a) Quadratic temperature–mortality fit for LRI without reference lines (b2 = 0.0292, p = 0.963; turning point 35.8 °C, far outside support); (b) observed temperature support 6.66–8.13 °C (shaded band) with the fitted curve; (c) between-zone versus within-zone variance decomposition of the four provincial thermal metrics (33 provinces × 30 years).

### 3.5 Falsification tests and climate-zone patterns

Figure 4 summarizes the falsification suite. The future-exposure test failed: mean-temperature coefficients at leads t + 1 and t + 2 were essentially identical to the concurrent estimates (LRI: −1.649 and −1.629 versus −1.696; asthma: −1.538 and −1.488 versus −1.600), meaning that future climate “predicts” current mortality as strongly as concurrent climate—the signature of trend-driven association rather than a causal lag structure. In 200 year permutations, the observed mean-temperature coefficient lay outside the permutation null distribution (0 of 200 permuted estimates reached the observed magnitude, p < 0.005; null SD ≈ 0.42), but this test only establishes that year alignment is non-random and cannot distinguish causation from shared trends. Together with the null detrended estimates (Section 3.3), these results indicate that the baseline associations reflect trend-level co-movement of declining mortality and changing climate; short-cycle causal effects are not identifiable in this national annual series.

**Figure 4.**
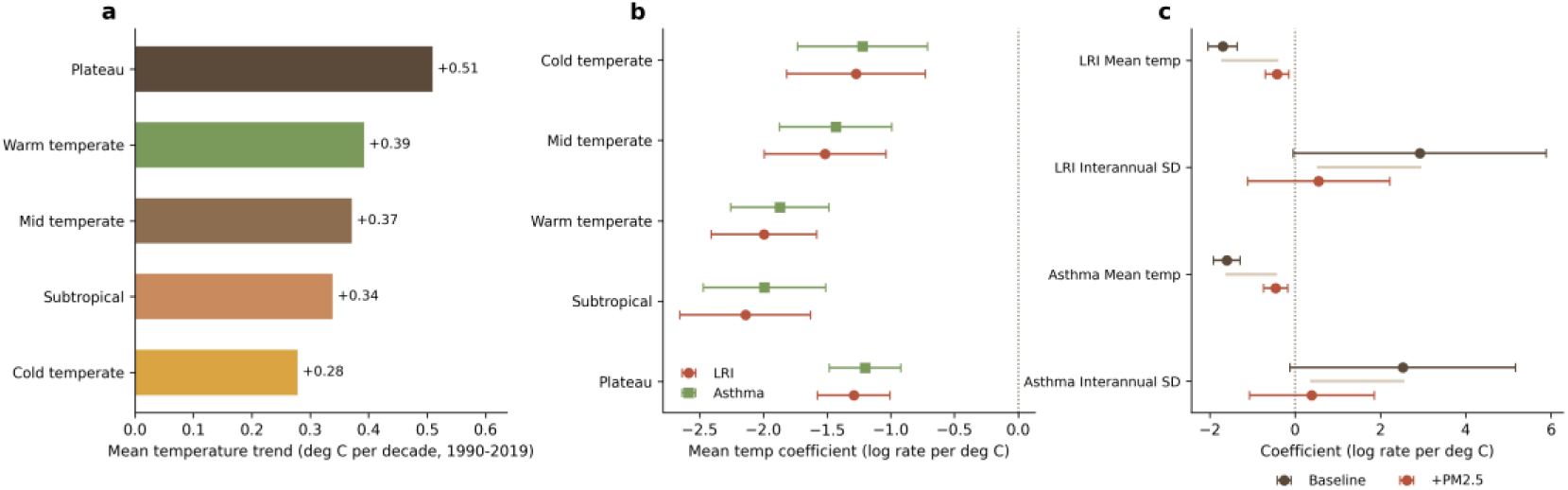
(a) Climate-zone warming trends, 1990–2019 (°C per decade, ranked); (b) zone-stratified mean-temperature coefficients (national LRI/asthma log rates versus zone climate; Newey–West lag 3); (c) baseline versus PM2.5-adjusted coefficients for mean temperature and interannual SD.

Figure 5 presents the provincial mean annual-temperature heatmap (33 provinces × 30 years, 1990–2019, provinces ordered by climate zone) and the sensitivity

**Figure 5.**
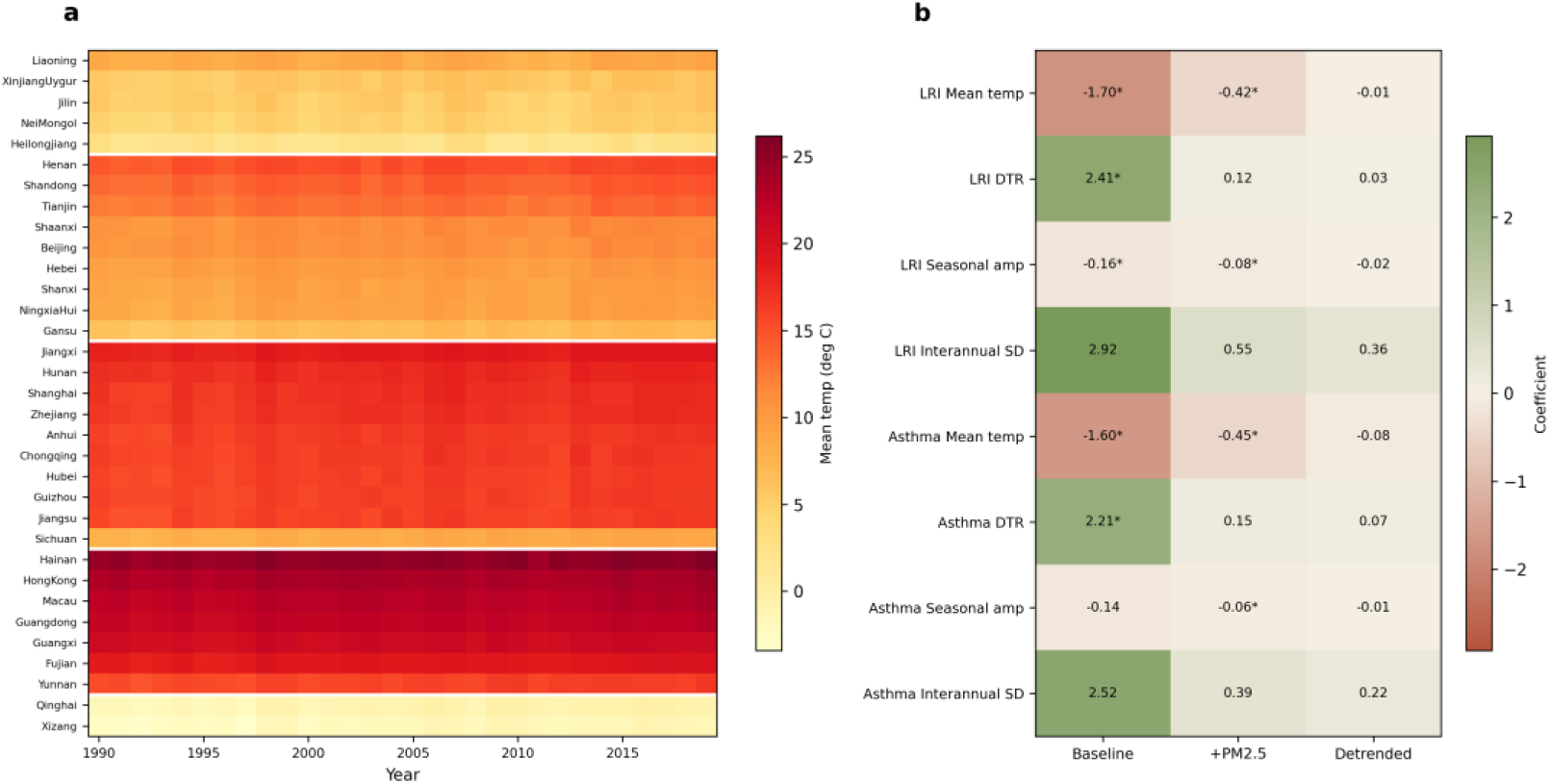
(a) Heatmap of provincial mean annual temperature, 33 provinces × 30 years (1990–2019), ordered by climate zone (white rules separate zones); (b) sensitivity summary: coefficients across baseline, PM2.5-adjusted and detrended specifications for all four thermal metrics (star: p < 0.05).

### 3.6 Projected changes in China’s thermal regime to the 2050s

China’s national mean temperature is projected to rise by +2.19 °C under SSP1-2.6, +2.46 °C under SSP2-4.5 and +3.02 °C under SSP5-8.5 (2041–2060 versus 1970–2000; Table 3). These cumulative changes span a ≈65-year interval (0.33–0.46 °C per decade), whereas the observed 1990–2019 trend was 0.364 °C per decade (fitted; corresponding to +1.06 °C over 2.9 decades, with a raw endpoint difference of +0.78 °C) — only SSP5-8.5 is clearly faster than the observed rate. The projected change in mean diurnal temperature range is small (−0.01, −0.14 and −0.07 °C), directionally consistent with the observed narrowing (−0.092 °C per decade), and projected seasonal amplitude is essentially unchanged (+0.36, −0.05 and +0.32 °C). After bias-correcting the WorldClim baseline to the C-LSAT 1990 anchor (a level offset of 0.89 °C: 7.26 versus 6.37 °C), China’s projected 2041–2060 national mean temperature is approximately 9.4–10.3 °C on the C-LSAT scale across SSP1-2.6 to SSP5-8.5 — still far below the minimum-mortality temperatures estimated in multi-country studies (14.7–19.8 °C), reinforcing the conclusion that China lies on the low-temperature side of the optimum; we note, however, that an annual-mean construct is not fully comparable with daily-scale MMT estimates, and the unidentifiability of the national MMT (Section 3.4) nonetheless precludes translating this exposure projection into a mortality projection.

**Table 3.** Projected change in China’s national thermal indicators, 2041–2060 versus 1970–2000 (WorldClim v2.1 baseline and CMIP6 MRI-ESM2-0 scenarios; matched-cell national means over 34,111 grid cells)

| **Indicator** | **SSP1-2.6** | **SSP2-4.5** | **SSP5-8.5** |
|---|---|---|---|
| Δ Annual mean temperature (°C) | +2.19 | +2.46 | +3.02 |
| Δ Mean diurnal temperature range (°C) | −0.01 | −0.14 | −0.07 |
| Δ Seasonal amplitude (°C) | +0.36 | −0.05 | +0.32 |
| Projected 2041–2060 mean temperature (°C) | 8.6 | 8.8 | 9.4 |

Note: the WorldClim baseline national mean (6.4 °C) is lower than the C-LSAT-based series used in this study (7.26 °C in 1990) because of differing periods, datasets and weighting; projected values are therefore compared as changes, not absolute levels.

## 4. Discussion

### 4.1 Principal findings

In this national time-series analysis of China over three decades, childhood mortality from lower respiratory infections (LRI) fell by 96.3% and asthma mortality by 94.9%, while the national mean temperature rose by 0.364 °C per decade and the diurnal temperature range (DTR) narrowed by 0.092 °C per decade. Baseline regressions yielded strong associations between all four climate indicators and mortality, but three lines of evidence—the failure of the future-exposure falsification test, the collapse of every coefficient after linear detrending, and the attenuation of most coefficients after adjustment for fine particulate matter (PM2.5)—indicate that these associations reflect shared secular trends rather than short-run causal effects. Finally, the national minimum-mortality temperature (MMT) was not identifiable: the observed support of annual mean temperature spans only 6.66–8.13 °C, and the nominal turning point of 35.84 °C is an extrapolation artifact.

### 4.2 The 96% decline and concurrent warming: concordant, not attributable

The near-elimination of childhood respiratory mortality in China is best understood as a development and health-systems phenomenon. The decline coincided with large expansions in healthcare access, nutrition, sanitation and immunization, including the period of routine diphtheria–tetanus–pertussis vaccination and the subsequent introduction of vaccines against respiratory pathogens; improvements in air quality further compressed the residual burden.^19^ ^16^ Warming unfolded in parallel as a background process, and the strong baseline coefficients we report quantify that parallelism rather than any protective effect of higher temperature. What this study adds is a falsification framework that converts the truism “correlation is not causation” into an auditable set of tests: the fact that future climate predicts current mortality as strongly as concurrent climate, and that detrending nullifies every coefficient, provides a reproducible basis for rejecting a causal reading. Because national climate–health assessments increasingly inform policy,^11^ we suggest that this test suite—future exposure, permutation, and detrending—should accompany any single-series national analysis before trend-level associations are interpreted, and we present it here as a discipline for climate–health inference in China rather than as a methodological novelty.

### 4.3 China lies on the low-temperature side of the optimum

Within the observed range, warming and declining mortality moved in the same direction, and as shown in Section 3.4, China’s area-weighted national mean temperature sits well below the minimum-mortality temperatures estimated in multi-country studies, so modest warming and declining mortality moved together within the observed range — though the MMT itself could not be identified in the observed range. This reading is also consistent with global multi-country evidence that cold-attributable mortality substantially exceeds heat-attributable mortality,^1^ ^2^ and with paediatric evidence that temperature predominantly affects infectious, including respiratory, outcomes in children.^9^ Two cautions are essential. First, the exposure construct differs: our exposure is the annual area-weighted mean temperature, not the daily temperature distributions underlying MMT estimates, so the comparison is directional rather than quantitative. Second, our finding that the national MMT is not identifiable—with only 1.47 °C of observed support and a quadratic term far from significance—is itself the important result for risk assessment: China’s future climate–health burden depends on where the mortality–temperature curve bends, and this analysis demonstrates that a 30-year national series cannot locate that bend. Estimating the Chinese childhood MMT will require distributed-lag designs on daily, subnational data,^18^ not further extrapolation of annual aggregates. Consistently, the bias-corrected CMIP6 scenario projection (Section 3.6; WorldClim baseline adjusted to the C-LSAT 1990 anchor, offset 0.89 °C) places China’s 2041–2060 national mean temperature at approximately 9.4–10.3 °C on the C-LSAT scale under SSP1-2.6 to SSP5-8.5 — still far below multi-country MMT estimates (14.7– 19.8 °C), though the annual-mean and daily-MMT constructs are not fully comparable — so even the SSP5-8.5 change projected for the 2050s (+3.0 °C relative to the 1970–2000 baseline) would not, on current evidence, carry the country across an identifiable optimum; whether the marginal effect of warming turns adverse remains an empirical question that these data cannot settle.

### 4.4 The hierarchy of variability associations

The variability indicators behaved differently from mean temperature, and differently from short-term city-level evidence. Time-series studies in Chinese cities consistently report acute harms from large DTR on respiratory mortality and childhood asthma morbidity,^8^ ^20^ ^10^ and multi-country work attributes a measurable mortality fraction to temperature variability.^5^ Our baseline DTR coefficient (+2.408 for LRI) appears directionally consistent with that literature, but the interpretation is constrained by an uncomfortable coincidence: in China, DTR declined over the study period while mortality also declined, so the positive coefficient encodes two falling trends rather than a short-run mechanism. After PM2.5 adjustment the DTR and interannual-variability coefficients lost significance, and after detrending all variability coefficients were null. The evidence therefore does not support a short-cycle causal effect of thermal variability on childhood respiratory mortality at the annual aggregate level; this does not contradict the acute city-level effects, which operate at daily lags that an annual national series cannot resolve.

### 4.5 Limitations

Several limitations qualify these findings. First, statistical power is inherently limited by 30 annual national observations, and the ecological design precludes inference to individuals. Second, mortality rates use a fixed GLOBOCAN 2022 population denominator, so rate trends partly reflect numerator changes under a static denominator; the asthma series additionally divides deaths at ages 0–24 years by the population aged 0–19 years, a numerator–denominator mismatch we retained for consistency and disclosed rather than corrected. Third, area weighting over-weights large western provinces relative to the population distribution of Chinese children. Fourth, PM2.5 and mean temperature are strongly collinear (r = 0.67), so the attenuated PM2.5-adjusted temperature coefficient should not be read as a causal decomposition. Fifth, the narrow observed temperature support precludes MMT identification. Sixth, mortality estimates derive from GBD 2021 while PM2.5 covariates derive from GBD 2023, and cross-version inconsistencies are possible. Finally, the five climate zones are an analytical partition, not an official climatological classification, and zone-level contrasts are descriptive. These constraints notwithstanding, the falsification-based interpretation—concordant trends, no identifiable short-run effect, no identifiable national optimum—rests on the internal logic of the tests rather than on the precision of any single coefficient. The scenario projection added in this version is subject to analogous constraints — a single climate model without ensemble spread, unweighted grid-cell national aggregation, and a WorldClim baseline that is not level-calibrated to the C-LSAT series — and is therefore read only as scenario-conditional change.

## 5. Conclusions

Over three decades, childhood lower respiratory infection mortality in China fell by 96.3% and asthma mortality by 94.9%, while the national mean temperature rose by 0.364 °C per decade. Strong baseline climate–mortality associations did not survive a pre-specified falsification suite: future exposure predicted current mortality as strongly as concurrent exposure, and every coefficient collapsed after detrending. The decline is therefore concordant with, but not attributable to, warming, and the national minimum mortality temperature is not identifiable from the narrow observed support (6.66–8.13 °C). The practical takeaway is twofold: trend-level national associations must not be read as causal effects, and China lies on the low-temperature side of the childhood optimum, so the direction of future warming impacts remains undetermined. Future work should exploit provincial-level GBD estimates for China if they are released publicly, monthly or daily distributed-lag designs,^18^ and CMIP6 scenario projections to locate the Chinese childhood optimum and quantify warming-attributable burdens.^11^ The scenario extension added in this version (Sections 2.8 and 3.6) takes a first step in that direction, projecting a further +2.2 to +3.0 °C of national warming by 2041–2060 (0.33–0.46 °C per decade over a ≈65-year interval; only SSP5-8.5 exceeds the observed 1990–2019 rate of 0.364 °C per decade) while leaving the mortality response undetermined.

## Declarations

### Ethics approval and consent to participate

Not applicable. The study used publicly available, de-identified, aggregated estimates and required no ethics approval or consent.

## Consent for publication

Not applicable.

### Availability of data and materials

Mortality estimates are available from the GBD 2021 Results Tool (gbd2021.healthdata.org, version 8016). The C-LSAT high-resolution land surface air temperature dataset and the C-LDTR diurnal temperature range dataset are available on figshare (DOI: 10.6084/m9.figshare.28255505 and DOI: 10.6084/m9.figshare.28255568). The population denominator was taken from GLOBOCAN 2022. The analysis code and the assembled analysis panel are available from the corresponding author on reasonable request. The WorldClim v2.1 baseline and CMIP6-downscaled scenario fields are publicly available (https://www.worldclim.org; https://geodata.ucdavis.edu/cmip6); the national projection values are given in Table 3 and accompany this article as a CSV file (chn_ssp2050_projection.csv); the raster-extraction script used to produce them is available from the corresponding author upon reasonable request.

Funding This work was supported by the Beijing Science and Technology Nova Program Interdisciplinary Project (20230484439) and the Beijing High-Level Public Health Technical Talents Project (2023-02-33). The funder had no role in study design, data collection, data analysis, data interpretation, or writing of the report.

## Presentation

This work has not been presented at any scientific meeting.

Disclosure The authors declare no conflicts of interest. AI tools were used for language editing and for assistance with code generation and modification; the authors take full responsibility for the content.

Authors’ contributions SC conceived the study, led the research design, and is the principal investigator responsible for the study idea and overall conduct. DL and HC curated the data and performed the analyses. SC supervised the study and is the corresponding author. All authors read and approved the final manuscript.

## Data Availability

Availability of data and materials Mortality estimates are available from the GBD 2021 Results Tool (gbd2021.healthdata.org, version 8016). The C-LSAT high-resolution land surface air temperature dataset and the C-LDTR diurnal temperature range dataset are available on figshare (DOI: 10.6084/m9.figshare.28255505 and DOI: 10.6084/m9.figshare.28255568). The population denominator was taken from GLOBOCAN 2022. The analysis code and the assembled analysis panel are available from the corresponding author on reasonable request. The WorldClim v2.1 baseline and CMIP6-downscaled scenario fields are publicly available (https://www.worldclim.org; https://geodata.ucdavis.edu/cmip6); the national projection values are given in Table 3 and accompany this preprint as a CSV file (chn_ssp2050_projection.csv); the raster-extraction script used to produce them is available from the corresponding author upon reasonable request.

https://vizhub.healthdata.org/gbd-results/

https://cds.climate.copernicus.eu/

## Acknowledgements

We thank the Institute for Health Metrics and Evaluation and the Global Burden of Disease collaborative network for making the mortality estimates publicly available, and the C-LSAT team at Sun Yat-sen University for developing and sharing the high-resolution temperature and diurnal temperature range datasets.

## Additional files

**Additional file 1: Supplementary Tables S1–S5.** Trend anchors and climate trends (S1); main model coefficients with PM2.5-adjusted and detrended specifications (S2); MMT bootstrap details (S3); falsification suite results (S4); climate-zone and province-level summaries (S5).

## References

1. Gasparrini A, Guo Y, Hashizume M, et al. Mortality risk attributable to high and low ambient temperature: a multicountry observational study. Lancet. 2015;386(9991):369–375. doi:10.1016/S0140-6736(14)62114-0.

2. Zhao Q, Guo Y, Ye T, et al. Global, regional, and national burden of mortality associated with non-optimal ambient temperatures from 2000 to 2019: a three-stage modelling study. Lancet Planet Health. 2021;5(7):e415–e425. doi:10.1016/S2542-5196(21)00081-4.

3. Guo Y, Gasparrini A, Armstrong B, et al. Global variation in the effects of ambient temperature on mortality: a systematic evaluation. Epidemiology. 2014;25(6):781–789. doi:10.1097/EDE.0000000000000165.

4. Tobías A, Armstrong B, Gasparrini A. Brief report: investigating uncertainty in the minimum mortality temperature: methods and application to 52 Spanish cities. Epidemiology. 2017;28(1):72–76. doi:10.1097/EDE.0000000000000567.

5. Guo Y, Gasparrini A, Armstrong BG, et al. Temperature variability and mortality: a multi-country study. Environ Health Perspect. 2016;124(10):1554–1559. doi:10.1289/EHP149.

6. Lee W, Bell ML, Gasparrini A, et al. Mortality burden of diurnal temperature range and its temporal changes: a multi-country study. Environ Int. 2018;110:123–130. doi:10.1016/j.envint.2017.10.018.

7. Cheng J, Xu Z, Zhu R, et al. Impact of diurnal temperature range on human health: a systematic review. Int J Biometeorol. 2014;58(9):2011–2024. doi:10.1007/s00484-014-0797-5.

8. Kan H, London SJ, Chen H, et al. Diurnal temperature range and daily mortality in Shanghai, China. Environ Res. 2007;103(3):424–431. doi:10.1016/j.envres.2006.11.009.

9. Xu Z, Etzel RA, Su H, Huang C, Guo Y, Tong S. Impact of ambient temperature on children’s health: a systematic review. Environ Res. 2012;117:120–131. doi:10.1016/j.envres.2012.07.002.

10. Xu Z, Huang C, Su H, Turner LR, Qiao Z, Tong S. Diurnal temperature range and childhood asthma: a time-series study. Environ Health. 2013;12:12. doi:10.1186/1476-069X-12-12.

11. Romanello M, Walawender M, Hsu SC, et al. The 2024 report of the Lancet Countdown on health and climate change. Lancet. 2024;404(10465):1847–1896. doi:10.1016/S0140-6736(24)01822-1.

12. Wu Y, Liu X, Gao L, et al. Short-term exposure to extreme temperature and outpatient visits for respiratory diseases among children in the northern city of China: a time-series study. BMC Public Health. 2024;24(1):341. doi:10.1186/s12889-024-17814-5.

13. Institute for Health Metrics and Evaluation (IHME). GBD Results Tool. Seattle, WA: IHME. https://vizhub.healthdata.org/gbd-results

14. GBD 2021 Causes of Death Collaborators. Global burden of 288 causes of death and life expectancy decomposition in 204 countries and territories and 811 subnational locations, 1990–2021: a systematic analysis for the Global Burden of Disease Study 2021. Lancet. 2024;403(10440):2100–2132. doi:10.1016/S0140-6736(24)00367-2.

15. Wei S, Li Q, Xu Q, Li Z, Zhang H, Lin J. Updates to C-LSAT 2.1 and the development of high-resolution land surface air temperature and diurnal temperature range datasets. Earth Syst Sci Data. 2025;17(10):4985–5005. doi:10.5194/essd-17-4985-2025.

16. GBD 2023 Disease and Injury and Risk Factor Collaborators. Burden of 375 diseases and injuries, risk-attributable burden of 88 risk factors, and healthy life expectancy in 204 countries and territories, 1990–2023. Lancet. 2025;406(10513):1873–1922. doi:10.1016/S0140-6736(25)01637-X.

17. GBD 2023 Demographics Collaborators. Global age-sex-specific all-cause mortality and life expectancy estimates for 204 countries and territories and 660 subnational locations, 1950–2023. Lancet. 2025;406(10513):1731–1810. doi:10.1016/S0140-6736(25)01330-3.

18. Gasparrini A, Armstrong B, Kenward MG. Distributed lag non-linear models. Stat Med. 2010;29(21):2224–2234. doi:10.1002/sim.3940.

19. GBD 2023 Causes of Death Collaborators. Global burden of 292 causes of death in 204 countries and territories and 660 subnational locations, 1990–2023. Lancet. 2025;406(10513):1811–1872. doi:10.1016/S0140-6736(25)01917-8.

20. Zhou X, Zhao A, Meng X, et al. Acute effects of diurnal temperature range on mortality in 8 Chinese cities. Sci Total Environ. 2014;493:92–97. doi:10.1016/j.scitotenv.2014.05.116.

21. Fick SE, Hijmans RJ. WorldClim 2: new 1-km spatial resolution climate surfaces for global land areas. Int J Climatol. 2017;37(12):4302–4315. doi:10.1002/joc.5086.

