## Supplementary_Materials for "Warming, thermal variability, and the 96% decline in childhood respiratory-infection mortality in China: a national time-series analysis of the Global Burden of Disease Study 2021 and the C-LSAT high-resolution climate dataset"

Scope: GBD 2021 China national estimates × C-LSAT HRv1; LRI ages 0–19 / asthma ages 0–24; deaths; Newey–West lag = 3; Supplementary Tables S1–S5.

#### Supplementary Table S1. Trend anchors: childhood respiratory mortality (1990–2021) and national climate indicators (1990–2019), China

| Panel A. Mortality anchors | 1990 | 2000 | 2010 | 2019 | 2021 |
|---|---|---|---|---|---|
| LRI deaths, ages 0–19, n (95% UI) | 330,194 (281,178–385,855) | 123,542 (109,176–138,292) | 32,559 (28,495–37,300) | 14,991 (12,410–17,764) | 12,098 (9,669–14,891) |
| LRI mortality rate (per 100,000) | 103.94 | 38.89 | 10.25 | 4.72 | 3.81 |
| Asthma deaths, ages 0–24, n (95% UI) | 3,287 (2,220–4,263) | 1,320 (1,060–1,549) | 441 (374–553) | 200 (153–266) | 167 (126–228) |
| Asthma mortality rate (per 100,000) | 1.035 | 0.416 | 0.139 | 0.063 | 0.052 |
| Panel B. National climate indicators | 1990 | 2019 | Slope per decade (°C) | | |
| Mean temperature (°C) | 7.26 | 8.04 | +0.364 | | |
| Diurnal temperature range (°C) | 12.09 | 12.01 | −0.092 | | |
| Seasonal amplitude (°C) | 27.01 | 27.34 | +0.250 | | |
| Interannual temperature SD (°C) | 0.443 | 0.444 | +0.0052 | | |

Notes. Panel A: deaths are GBD 2021 estimates with 95% uncertainty intervals (UI, lower–upper) in parentheses; rates use the fixed GLOBOCAN 2022 denominator of the Chinese population aged 0–19 years (317,671,474). Between 1990 and 2021, LRI deaths fell by 96.3% and asthma deaths by 94.9%. Panel B: climate indicators are land-area-weighted national aggregates of the C-LSAT HRv1 / C-LDTR 0.5° grids (1990–2019); slopes are ordinary least squares per-decade trends over 1990–2019 (per 2021 where the series ends in 2019). Cells left blank in Panel B are not applicable.

#### Supplementary Table S2. Main model coefficients: log mortality rate per +1 °C, Newey–West HAC (lag = 3), n = 30 (1990–2019)

| Outcome | Specification | Indicator | β | SE | 95% CI | p | n |
|---|---|---|---|---|---|---|---|
| LRI | Baseline | Mean temperature | −1.696 | 0.174 | (−2.037–−1.355) | <0.001 | 30 |
| LRI | Baseline | Diurnal temperature range | 2.408 | 0.336 | (1.750–3.066) | <0.001 | 30 |
| LRI | Baseline | Seasonal amplitude | −0.162 | 0.082 | (−0.322–−0.002) | 0.047 | 30 |
| LRI | Baseline | Interannual temperature SD | 2.924 | 1.514 | (−0.044–5.892) | 0.054 | 30 |
| LRI | +PM2.5 SEV | Mean | −0.419 | 0.140 | (−0.692– | 0.003 | 30 |

| Outcome | Specification | Indicator | β | SE | 95% CI | p | n |
|---|---|---|---|---|---|---|---|
| | | temperature | | | −0.145) | | |
| LRI | +PM2.5 SEV | Diurnal temperature range | 0.117 | 0.337 | (−0.543–0.777) | 0.728 | 30 |
| LRI | +PM2.5 SEV | Seasonal amplitude | −0.077 | 0.032 | (−0.140–−0.015) | 0.015 | 30 |
| LRI | +PM2.5 SEV | Interannual temperature SD | 0.553 | 0.850 | (−1.112–2.218) | 0.515 | 30 |
| LRI | Detrended | Mean temperature | −0.010 | 0.068 | (−0.144–0.124) | 0.882 | 30 |
| LRI | Detrended | Diurnal temperature range | 0.031 | 0.123 | (−0.211–0.273) | 0.802 | 30 |
| LRI | Detrended | Seasonal amplitude | −0.016 | 0.014 | (−0.043–0.012) | 0.271 | 30 |
| LRI | Detrended | Interannual temperature SD | 0.357 | 0.583 | (−0.785–1.500) | 0.540 | 30 |
| Asthma | Baseline | Mean temperature | −1.600 | 0.161 | (−1.916–−1.284) | <0.001 | 30 |
| Asthma | Baseline | Diurnal temperature range | 2.210 | 0.294 | (1.634–2.787) | <0.001 | 30 |
| Asthma | Baseline | Seasonal amplitude | −0.137 | 0.072 | (−0.278–0.003) | 0.055 | 30 |
| Asthma | Baseline | Interannual temperature SD | 2.525 | 1.348 | (−0.117–5.167) | 0.061 | 30 |
| Asthma | +PM2.5 SEV | Mean temperature | −0.453 | 0.145 | (−0.738–−0.168) | 0.002 | 30 |
| Asthma | +PM2.5 SEV | Diurnal temperature range | 0.152 | 0.310 | (−0.456–0.760) | 0.625 | 30 |
| Asthma | +PM2.5 SEV | Seasonal amplitude | −0.061 | 0.029 | (−0.118–−0.004) | 0.037 | 30 |
| Asthma | +PM2.5 SEV | Interannual temperature SD | 0.395 | 0.747 | (−1.070–1.859) | 0.597 | 30 |
| Asthma | Detrended | Mean temperature | −0.084 | 0.072 | (−0.225–0.058) | 0.247 | 30 |
| Asthma | Detrended | Diurnal temperature range | 0.071 | 0.077 | (−0.079–0.222) | 0.353 | 30 |
| Asthma | Detrended | Seasonal amplitude | −0.005 | 0.011 | (−0.027–0.016) | 0.631 | 30 |
| Asthma | Detrended | Interannual temperature SD | 0.215 | 0.401 | (−0.571–1.002) | 0.591 | 30 |

Notes. Ordinary least squares of the natural logarithm of the national mortality rate on the four climate indicators jointly, n = 30 annual observations (1990–2019); standard errors (SE) are Newey–West HAC with a maximum lag of 3 years. 95% CI = 95% confidence interval (lower–upper). Specifications: Baseline (four indicators only); +PM2.5 SEV (adds the GBD 2023 national ambient PM2.5 summary exposure value as a covariate); Detrended (log rate and all regressors residualised on calendar year). The PM2.5 SEV coefficient itself is reported in Supplementary Table S4 (Panel D).

### Supplementary Table S3. Minimum mortality temperature (MMT): quadratic-model and bootstrap details

| Outcome | b1 (linear) | b2 (quadratic) | p (b2) | Nominal turning point (°C) | Bootstrap 95% CI (°C) | Valid resamples (of 500) | Observed support (°C) | Identifiability |
|---|---|---|---|---|---|---|---|---|
| LRI | −2.093 | 0.029 | 0.963 | 35.84 | 7.92–46.50 | 248 / 500 | 6.66–8.13 | Not identifiable: turning point outside observed support (extrapolation artifact) |
| Asthma | −0.900 | −0.045 | 0.935 | None (b2 ≤ 0) | 8.03–45.10 | 218 / 500 | 6.66–8.13 | Not identifiable: no interior minimum |

Notes. A quadratic in annual mean temperature (log rate on mean_temp and mean_temp²) was fitted on n = 30 years (1990–2019); the turning point is −b1/2b2. Uncertainty used 500 bootstrap resamples over years; draws with a non-positive quadratic coefficient (no interior minimum) were discarded, and the count retained is shown. “Not identifiable” is declared when (i) the quadratic term is non-significant or non-positive, and/or (ii) the nominal turning point lies outside the observed exposure support. For LRI the nominal turning point of 35.84 °C lies far outside the observed support of 6.66–8.13 °C and is an extrapolation artifact; for asthma the quadratic term is negative, so no interior minimum exists. The national childhood MMT is therefore not identifiable from this series.

### Supplementary Table S4. Falsification suite: future-exposure leads, year permutation, detrended specification, and the PM2.5 coefficient

| Panel A. Concurrent vs future exposure (t + 1, t + 2) | Indicator | β | p |
|---|---|---|---|
| LRI, Concurrent (t) | Mean temperature | −1.696 | <0.001 |
| LRI, Concurrent (t) | Diurnal temperature range | 2.408 | <0.001 |
| LRI, Concurrent (t) | Seasonal amplitude | −0.162 | 0.047 |
| LRI, Concurrent (t) | Interannual temperature SD | 2.924 | 0.054 |
| LRI, Future (t + 1) | Mean temperature | −1.649 | <0.001 |
| LRI, Future (t + 1) | Diurnal temperature range | 2.302 | <0.001 |
| LRI, Future (t + 1) | Seasonal amplitude | −0.175 | 0.022 |
| LRI, Future (t + 1) | Interannual temperature SD | 2.951 | 0.055 |
| LRI, Future (t + 2) | Mean temperature | −1.629 | <0.001 |
| LRI, Future (t + 2) | Diurnal temperature range | 2.184 | <0.001 |
| LRI, Future (t + 2) | Seasonal amplitude | −0.164 | 0.043 |
| LRI, Future (t + 2) | Interannual temperature SD | 2.865 | 0.069 |
| Asthma, Concurrent (t) | Mean temperature | −1.600 | <0.001 |
| Asthma, Concurrent (t) | Diurnal temperature range | 2.210 | <0.001 |
| Asthma, Concurrent (t) | Seasonal amplitude | −0.137 | 0.055 |
| Asthma, Concurrent (t) | Interannual temperature SD | 2.525 | 0.061 |
| Asthma, Future (t + 1) | Mean temperature | −1.538 | <0.001 |
| Asthma, Future (t + 1) | Diurnal temperature range | 2.132 | <0.001 |
| Asthma, Future (t + 1) | Seasonal amplitude | −0.153 | 0.034 |
| Asthma, Future (t + 1) | Interannual temperature SD | 2.442 | 0.071 |
| Asthma, Future (t + 2) | Mean temperature | −1.488 | <0.001 |
| Asthma, Future (t + 2) | Diurnal temperature range | 2.027 | <0.001 |
| Asthma, Future (t + 2) | Seasonal amplitude | −0.145 | 0.063 |
| Asthma, Future (t + 2) | Interannual temperature SD | 2.276 | 0.108 |

Notes. Panel A: specifications identical to the baseline model but with exposures replaced by their t + 1 and t + 2 leads (n = 29 and 28 years, respectively). A genuinely causal concurrent effect should attenuate at future leads; instead, future-exposure coefficients are essentially identical to the concurrent estimates, so the future-exposure test fails.

| Panel B. Year permutation (200 draws), mean-temperature coefficient | Observed β | Permutation mean | Permutation SD | Permutation 95% range | Permutation p |
|---|---|---|---|---|---|
| LRI | −1.696 | 0.018 | 0.423 | −0.772–0.765 | <0.005 (0/200) |
| Asthma | −1.600 | 0.058 | 0.402 | −0.732–0.835 | <0.005 (0/200) |

Notes. Panel B: calendar years of the mortality series were permuted 200 times; no permuted estimate reached the observed magnitude. This test establishes that year alignment is non-random but cannot distinguish causation from shared trends.

| Panel C. Detrended specification | Indicator | β | SE | p |
|---|---|---|---|---|
| LRI | Mean temperature | −0.010 | 0.068 | 0.882 |
| LRI | Diurnal temperature range | 0.031 | 0.123 | 0.802 |
| LRI | Seasonal amplitude | −0.016 | 0.014 | 0.271 |
| LRI | Interannual temperature SD | 0.357 | 0.583 | 0.540 |
| Asthma | Mean temperature | −0.084 | 0.072 | 0.247 |
| Asthma | Diurnal temperature range | 0.071 | 0.077 | 0.353 |
| Asthma | Seasonal amplitude | −0.005 | 0.011 | 0.631 |
| Asthma | Interannual temperature SD | 0.215 | 0.401 | 0.591 |

Notes. Panel C: log rate and all regressors linearly detrended on calendar year (n = 30). Every climate coefficient collapses toward zero and none approaches significance (all p > 0.24), indicating that the baseline associations are trend-level co-movement.

| Panel D. PM2.5 summary exposure value, own coefficient | β | SE | 95% CI | p |
|---|---|---|---|---|
| LRI | −0.070 | 0.008 | (−0.086–−0.055) | <0.001 |
| Asthma | −0.063 | 0.008 | (−0.078–−0.048) | <0.001 |

Notes. Panel D: coefficient on the GBD 2023 national ambient PM2.5 summary exposure value in the PM2.5-adjusted specification. The negative coefficient co-moves with the long-run mortality decline and should not be read as a short-run protective effect of PM2.5; PM2.5 and mean temperature are strongly collinear (r = 0.67).

### Supplementary Table S5. Climate-zone and province-level summaries of the four climate indicators, 1990–2019

| Panel A. Climate-zone means | Mean temperature (°C) | DTR (°C) | Seasonal amplitude (°C) | Interannual SD (°C) |
|---|---|---|---|---|
| Cold temperate (NE/N Xinjiang) | 5.01 ± 0.47 | 13.10 ± 0.25 | 35.91 ± 1.56 | 0.484 ± 0.063 |
| Mid temperate (N & NW China) | 10.16 ± 0.50 | 11.84 ± 0.27 | 27.97 ± 1.27 | 0.436 ± 0.067 |
| Plateau (Qinghai-Tibet) | −1.67 ± 0.58 | 14.37 ± 0.29 | 23.61 ± 1.00 | 0.394 ± 0.085 |
| Subtropical (S China) | 18.94 ± 0.40 | 9.10 ± 0.23 | 14.63 ± 1.01 | 0.347 ± 0.040 |
| Warm temperate (Yangtze basin) | 14.35 ± 0.46 | 9.17 ± 0.25 | 21.76 ± 1.23 | 0.365 ± 0.043 |

Notes. Panel A: entries are the across-year mean ± standard deviation of each area-weighted zone aggregate over 1990–2019. Zones: cold temperate (Heilongjiang, Jilin, Liaoning, NeiMongol, XinjiangUygur); mid temperate (Beijing, Tianjin, Hebei, Shanxi, Shandong, Henan, Shaanxi, Gansu, NingxiaHui); warm temperate (Shanghai, Jiangsu, Anhui, Zhejiang, Hubei, Hunan, Jiangxi, Sichuan, Chongqing, Guizhou); subtropical (Fujian, Guangdong, Guangxi, Hainan, Yunnan, HongKong, Macau); plateau (Qinghai, Xizang).

| Panel B. Climate-zone trends, °C per decade (1990–2019) | Mean temperature | DTR | Seasonal amplitude | Interannual SD |
|---|---|---|---|---|
| Cold temperate (NE/N Xinjiang) | +0.278 | −0.104 | +0.419 | +0.0148 |
| Mid temperate (N & NW China) | +0.371 | −0.108 | +0.195 | −0.0027 |
| Plateau (Qinghai-Tibet) | +0.509 | −0.196 | +0.117 | −0.0039 |
| Subtropical (S China) | +0.339 | −0.018 | +0.122 | −0.0045 |
| Warm temperate (Yangtze basin) | +0.392 | +0.018 | +0.152 | +0.0065 |

Notes. Panel B: ordinary least squares per-decade slopes of each zone aggregate. The plateau zone warmed fastest (+0.509 °C per decade). Zone-level contrasts are descriptive and support no inference.

| Panel C. Province-level means, 1990–2019 | Mean temperature (°C) | DTR (°C) | Seasonal amplitude (°C) | Interannual SD (°C) |
|---|---|---|---|---|
| Anhui | 16.39 | 8.90 | 25.33 | 0.388 |
| Beijing | 11.00 | 11.76 | 30.89 | 0.436 |
| Chongqing | 16.30 | 7.34 | 21.61 | 0.397 |
| Fujian | 19.08 | 8.55 | 17.58 | 0.334 |
| Gansu | 6.62 | 13.33 | 27.49 | 0.408 |
| Guangdong | 21.94 | 7.93 | 15.58 | 0.352 |
| Guangxi | 20.90 | 7.76 | 16.95 | 0.393 |
| Guizhou | 16.03 | 7.71 | 19.39 | 0.389 |
| Hainan | 25.03 | 7.47 | 10.12 | 0.381 |
| Hebei | 9.90 | 11.97 | 31.26 | 0.446 |
| Heilongjiang | 2.26 | 12.11 | 41.98 | 0.592 |
| Henan | 15.08 | 10.05 | 26.28 | 0.420 |
| HongKong | 23.43 | 6.77 | 14.10 | 0.329 |
| Hubei | 16.07 | 8.45 | 23.76 | 0.388 |
| Hunan | 17.62 | 7.90 | 22.92 | 0.363 |
| Jiangsu | 15.88 | 8.40 | 25.70 | 0.422 |
| Jiangxi | 18.66 | 8.39 | 22.30 | 0.331 |
| Jilin | 5.21 | 11.93 | 38.31 | 0.595 |
| Liaoning | 8.46 | 11.18 | 34.31 | 0.545 |
| Macau | 22.60 | 5.88 | 14.00 | 0.373 |
| NeiMongol | 4.65 | 13.31 | 37.37 | 0.494 |
| NingxiaHui | 9.11 | 12.56 | 28.76 | 0.490 |
| Qinghai | −1.15 | 14.32 | 24.35 | 0.403 |
| Shaanxi | 11.18 | 11.19 | 25.97 | 0.442 |
| Shandong | 13.99 | 9.81 | 27.88 | 0.433 |
| Shanghai | 17.13 | 7.29 | 24.62 | 0.438 |
| Shanxi | 9.34 | 12.40 | 29.02 | 0.489 |
| Sichuan | 8.32 | 11.56 | 19.02 | 0.329 |
| Tianjin | 13.09 | 10.83 | 30.54 | 0.430 |
| XinjiangUygur | 5.70 | 13.53 | 33.06 | 0.428 |
| Xizang | −2.01 | 14.40 | 23.15 | 0.388 |
| Yunnan | 15.75 | 10.80 | 12.21 | 0.317 |
| Zhejiang | 17.01 | 8.40 | 23.07 | 0.390 |

Notes. Panel C: across-year means for the 33 province-level units (GADM 4.1 boundaries), aggregated from the C-LSAT HRv1 / C-LDTR 0.5° grids with land-area weights. Provincial paediatric population time series were unavailable, so province values are descriptive climate summaries only.

*End of Supplementary Materials.*
